# Generalisability of epileptiform patterns across time and patients

**DOI:** 10.1101/2023.08.29.23294708

**Authors:** Hamid Karimi-Rouzbahani, Aileen McGonigal

## Abstract

Complexity of epileptogenic zone (EZ) localisation contributes to failure of surgical resection to produce seizure freedom. This is to some extent a result of distinct patterns of epileptiform activity between (i.e., interictal) and during seizures (i.e., ictal) and their diversity across patients. This often leads to suboptimal localisation based on inspection of electroencephalography (EEG) features. We asked two open questions. First, whether neural signal reflecting epileptogenicity would be generalisable from interictal to ictal time window within each patient. This would be critical for patients who are monitored in hospital without having a seizure to help with EZ localisation, and more generally for understanding the predictive power of resting state (interictal) EEG data in determining EZ. Second, whether epileptiform patterns would generalise across patients, and if so, which aspects of those patterns are the most generalisable.

We used an intracranial EEG dataset that included fifty-five patients with lesional and non-lesional pathology, who had subsequently undergone cortical resection in frontal or temporal lobe with different levels of seizure freedom. We extracted a large set of simple to complex features from stereo-EEG (SEEG) and electrocorticographic (ECoG) neural signals recorded during interictal and ictal time windows. We fed those features to decision tree classifiers for EZ localisation and to quantify the diversity of ictal and interictal epileptiform patterns through a cross-time and cross-patient generalisation procedure.

We observed significant evidence (*Bayes factor* >> 10) for generalisability of patterns from interictal to ictal time windows across patients, which were dominantly reflected in signal power and high-frequency network-based features. Majority of patients showed consistent patterns of epileptogenicity across interictal and ictal time windows, reflected in above-chance area-under-curve (mean *AUC =* 0.6). We observed significant evidence (*Bayes factor* >> 10) that signal features of epileptogenic regions could generalise across patients in both interictal and ictal time windows with significant evidence for higher generalisability in ictal than interictal time window (mean *AUC* 0.75 vs. 0.59; *Bayes factor* >> 10). While signal power and moment features were the most contributory to the cross-patient generalisation in the interictal window, signal complexity features were the most contributory in the ictal window.

These results provide new insights about features of epileptic neural activity that generalise across interictal-ictal time windows and patients, which can have implications for both qualitative and quantitative EZ localisation. The explainable machine-learning pipeline developed here can guide future developments in epilepsy investigations.

## Introduction

There are over 50 million people with epilepsy worldwide^1^. Anti-seizure medications cannot adequately control the disorder in about 30% of cases^2^. If the epilepsy is considered focal (i.e., seizures arising from part of one hemisphere^3^), those with drug-resistant focal epilepsy may undergo presurgical evaluation to detect areas involved in the generation of seizure activity, which may require intracranial electroencephalography (EEG) in some. These areas can be collectively referred to as the epileptogenic zone (EZ), a term that was conceptually developed from stereo-electroencephalography (SEEG)^4^, a method of intracerebral recording based on multiple depth electrodes. The EZ is considered as the region of primary seizure organization^5^. After localisation, if the clinical risk-benefit ratio is deemed favourable for a specific patient, the EZ can be removed and/or disconnected through surgical resection or laser-based ablation. Despite great progress in use of multimodal approaches (e.g., magnetic resonance imaging (MRI), electroencephalography (EEG), positron emission tomography (PET) scans, etc.) and wealth of clinical expertise, precise localisation of the EZ often remains difficult and may lead to failure to achieve seizure freedom^6,7^.

Quantification methods have shown great promise in localising the EZ through quantification of intracranial EEG signals^8–10(see11^ for review). These methods generally investigate either the interictal or the ictal time window (see Supplementary Table 1 for an overview). In the ictal window, low voltage fast activity (LVFA), baseline shift, rhythmic spikes/spike-waves and preictal low frequency spiking, were found to be the most prevalent epileptiform activities^12^. These characteristics were successfully extracted from signals and used for EZ localisation in several studies^13–19^. In the interictal window, the traditional epileptiform characteristics include interictal spikes/discharges^20^ and high-frequency oscillations (HFOs^21^) with a debate on more efficacy of one over the other, and ultimately possible increased predictive EZ by measuring their co-occurrence^22^. Modelling approaches based on patient-specific imaging data can predict spatial extent of epileptogenicity^23^.

The relation between electrical seizure onset and electrical disturbances detectable between seizures is of great clinical and neuroscientific importance and yet remains incompletely known^20^. Interictal spiking is a heterogeneous phenomenon that reflects the involvement of different neuronal networks and mechanisms (e.g., synaptic conductance) in different regions of an epileptic brain^24^ and has shown predictive value in differentiating underlying neuropathological substrates^25^. In fact, the original concept of “epileptic focus” arose not from seizure recordings, but from observations of interictal spiking during ECoG^5^. Basing surgical excision extent on the zone of interictal spiking measured peri-operatively by ECoG was thought to help improve likelihood of surgical outcome, albeit with variable accuracy^26^. In pioneering SEEG work^27^, the regions involved by interictal spiking typically showed (partial) overlap with the zone of primary seizure organization. Observations from clinical data of this type as well as animal models suggest that interictal and ictal signal features may reflect some similar neuronal mechanisms^24^. Apart from spikes and HFOs, many other linear and nonlinear signal features including randomness, power and entropy have shown success in localising EZ in the interictal window^28–35^. While many of the classical methods for EZ localisation relied on univariate/single-channel signal activity, there has been a shift to multivariate/network-based localisation^10,35–42^, which aligns with the conceptualisation of epilepsy as a network disorder^43–45^ and has shown better performance than univariate methods^10,38,42,46–48^.

Despite the large literature on EZ localisation, using various signal analysis approaches applied to both interictal and ictal windows (Supplementary Table 1), the correspondence between the two windows has remained unclear. This might be because of the distinct, pre-defined sets of features which were looked for in the two windows and which appear visually different (e.g., interictal spikes and ictal LVFA). Nonetheless, if there are signal features which are shared between the two windows, interictal activity, which represents most of the patient’s brain state and is generally easier to record, could potentially be sufficient to predict EZ. As a first step to explore this, we looked at a heterogeneous group of epilepsies studied with intracranial EEG, available in an open-access dataset^46,48^. We compared a large battery of explainable signal characteristics, ranging from simple single-channel to computationally complex network-based features, from both interictal and ictal time windows, to see which features generalise across the two time-windows using the data from each individual patient (i.e., within-individual across-time generalisation^1^). Moreover, to see if there are features which are shared between individuals, we also evaluated the generalisability of features across patients within each individual time window (i.e., across-individual within-time generalisation). Finally, we evaluated the effect of surgical outcome (seizure-free/not seizure-free), EZ, pathology of epilepsy (lesional/non-lesional) and type of recording (SEEG/ECoG) on the EZ localisation performances.

## Materials and methods

### Dataset

This study uses a well-structured open-access intracranial dataset which brings together data from multiple centres^46,48^. The dataset includes 57 patients who had been implanted with either subdural grid/strip (termed “electrocorticography” (ECoG)^47^) or SEEG as their presurgical workup, and subsequently treated with surgical resection or laser ablation. Clinically determined seizure onset channels were provided, as well as marking of channels which overlap with the resection/ablation zone, which was rigorously determined by segmenting the resection cavity. Two patients’ data were excluded from our analyses as one had no interictal and the other no ictal recordings. Each patient had 2 interictal recordings and between 1 to 5 (*mean =* 3.7) ictal recordings/seizures (110 interictal and 204 ictal recordings over all patients). The interictal data was selected from awake brain activities determined both by the selection of day-time epochs (8 am – 8 pm) and the use of a custom non-REM sleep detector (explained in detail in Bernabei et al.,^48^). The interictal data were at least 2 hours before the beginning of a seizure and at least 2 hours after a subclinical seizure, 6 hours after a focal seizure and 12 hours after a generalised seizure. The sampling frequencies of the signals varied across patients and ranged from 256 to 1024 Hz. We adjusted the sampling rate of all datasets to 256 Hz across patients. The details of the patients included in the analyses are provided in Supplementary Table 2. Epileptogenic zones/resected areas ranged from frontal, frontoparietal, mesiofrontal, temporal, mesiotemporal, parietal and insular areas.

### Pre-processing

We used a 5-minute signal from each interictal recording (10 minutes per patient) and a 58-second signal from each ictal recording (−30 to +28 seconds around the time of seizure onset). Bad channels, as marked in the dataset, were excluded from analyses. An average of 105.6 contacts (*std* = 38.04) per patient remained for analysis after bad channels were removed from the dataset. There was an average of 114.2 (*std* = 41.2) and 88.8 (*std* = 25.3) channels recorded in patients implanted with SEEG and ECoG, respectively. Among these, an average of 12.87% (*std* = 11.1%) of channels were in the EZ/resected area in each patient. We applied no filtering or artefact removal on the dataset. As the low- and high-frequency noise is shared across both groups of contacts, and as classifiers rely on the differences between classes rather than similarities^49^, we did not apply filters. Moreover, by not applying any filters, we allowed easier replication of results in future studies as any choice of filters can potentially affect the results in some way^50^.

### Feature extraction

We quantified the signal patterns by extracting 34 mathematically distinct features. Features were extracted in 2s non-overlapping sliding windows along the interictal and ictal signals as in previous studies^51–54^. This led to 14 pre- and 14 post-seizure onset time windows in the ictal period excluding the last window. To quantify changes to neural activities upon seizure onset, we normalised the extracted post-seizure onset data by the pre-seizure onset data using equation (1):

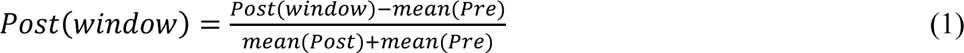

where *Pre* and *Post* refer to the arrays of 14 feature values extracted from neural data. This led to 14 normalised ictal feature values which were used for analysis. In interictal data, we down sampled the number of extracted feature samples (n ∼ 150) to 28 samples using the Matlab “resample” function. This led to approximately equal number of data points in interictal and ictal windows, from 110 interictal and 204 ictal recordings. A range of simple to complex signal features were extracted. All these features have been previously used to quantify EEG patterns^55–57^ and the reader is referred to Supplementary Text 1 and the mentioned publications for details. Briefly, we extracted four categories of signal features to obtain a relatively comprehensive view of signal characteristics. These include the signal *moment* features, nonlinear *complexity* features, *frequency*-domain features and *network*-based features.

### Multivariate pattern classification

We used a standard multivariate pattern classification procedure to localise EZ (i.e., to discriminate epileptogenic/resected and non-epileptogenic/non-resected contacts). We use the term “epileptiform” patterns/activities in a general sense to refer to any patterns which discriminated epileptogenic and non-epileptogenic contacts. Accordingly, the classification performance indicates how discriminable were the signal patterns across these two sets of contacts. We quantified the classification performance by area-under-the-curve (*AUC*) to provide a comprehensive, threshold-free classification performance^49^. As in recent studies^48,58^, we used decision tree (DT) classifiers, and each contact was treated like an observation in classification. Our DT classifiers used a random forest algorithm with 50 bags of feature combinations. DT classifiers are well suited for nonlinear feature classifications and provide insights into feature contributions. This method also provides a “feature contribution” metric by permuting the observation/contact labels in each feature separately and quantifying its effect on performance-contribution is in inverse proportion to performance drop. We performed three distinct types of classifications - one within patient and time (non-generalisation) and two which involved generalisation either across time or patients. In all three analyses, we classified epileptogenic and non-epileptogenic contacts (i.e., EZ localisation). In the non-generalisation classification, we performed the classification within the interictal and ictal time window separately for each patient using a 10-fold cross-validation procedure. In the cross-time generalisation, in each patient, we trained the classifier using the data from the two interictal recordings and tested the classifier using all the ictal recordings (*mean =* 3.7). In the cross-patient generalisation, in each time window (interictal/ictal), we trained the classifier using the data from all patients minus one and tested the classifier using the data from the left-out patient and repeated this procedure until every patient was used once for testing the classifier.

To equalise the number of epileptogenic to non-epileptogenic contacts (12.87% vs. 77.13% on average, respectively) which is essential for avoiding bias toward one class in classification^49^, we used a down-sampling procedure in our analyses and repeated every classification of data 1000 times before averaging the results.

To generate chance-level performances, against which we could evaluate the validity of our true classification performances, we shuffled (epileptogenic/non-epileptogenic) contact labels 1000 times and recalculated the classification performance leading to 1000 chance-level classification results.

### Statistical analysis

We used a Bayes Factor analysis for statistical inference^59^. We compared the levels of *AUCs* against chance-level *AUCs* as well as evaluated main effects on classifications. We used standard rules of thumb for interpreting levels of evidence^60,61^: Bayes factors between 3 and 10 and between 1/10 and 1/3 were interpreted as evidence for the alternative and null hypotheses, respectively. Bayes factors > 10 and < 1/10 were interpreted as significant evidence for the alternative and null hypotheses, respectively. We considered the Bayes factors which fell between 1/3 and 3 as insufficient evidence either way. Insufficient evidence means that no conclusions can be made about difference between a pair of variables.

To evaluate the evidence for the null and alternative hypotheses of at-chance and above-chance classification, respectively, we compared the classification rates in each analysis and the classification rates obtained from the chance-level classification results (e.g., panel A in all figures). For that, we performed an unpaired Bayes factor *t-test* for alternative (i.e., difference from chance; H1) and the null (i.e., no difference from chance; H0) hypotheses. To evaluate the evidence for the null and alternative hypotheses of difference between classification levels across analyses (e.g., Interictal vs. Ictal), we compared the classification rates obtained from each of those analyses using paired Bayes factor *t-test*. To evaluate the main effects of resection outcome, EZ, pathology (lesional/non-lesional) and type of recording (SEEG/ECoG), we used a Bayes factor *ANOVA*, with these four factors as independent variables and classification/generalisation *AUC* as the dependent variable. For statistical power in *ANOVA*, we excluded patients with insular, frontoparietal, parietal and mesiofrontal resection which were under-sampled (n < 3). The priors for all Bayes factor analyses were determined based on Jeffrey-Zellner-Siow priors^62,63^ which are from the Cauchy distribution based on the effect size that is initially calculated in the algorithm using *t-test*^59^.

### Data and code availability

The dataset used in this study was from previous studies and is available at https://openneuro.org/datasets/ds004100/versions/1.1.3. The code developed for this project is available at https://github.com/HamidKarimi-Rouzbahani/Intracranial_EEG_generalisation.

## Results

We used a multivariate pattern analysis approach on features extracted from intracranial SEEG/ECoG data in patients with epilepsy to address two main questions. First, we wondered if there were similarities between the epileptiform patterns which discriminated epileptogenic from non-epileptogenic areas in interictal and ictal time windows. Second, we wondered how generalisable epileptiform patterns were across patients.

### How discriminable are epileptogenic and non-epileptogenic contacts?

As an initial step in our analyses, we quantified the discriminability of epileptogenic and non-epileptogenic contacts. This was done for each patient and time window (interictal and ictal) separately. There was significant evidence (*BF* >> 10) for above-chance *AUC* which averaged to 0.97 (*std* = 0.03) in the interictal and 0.91 (*std* = 0.07) in ictal time windows, respectively (Fig. 1A). These showed that our multi-feature classification pipeline could robustly differentiate epileptogenic from non-epileptogenic contacts.

**Figure 1.**
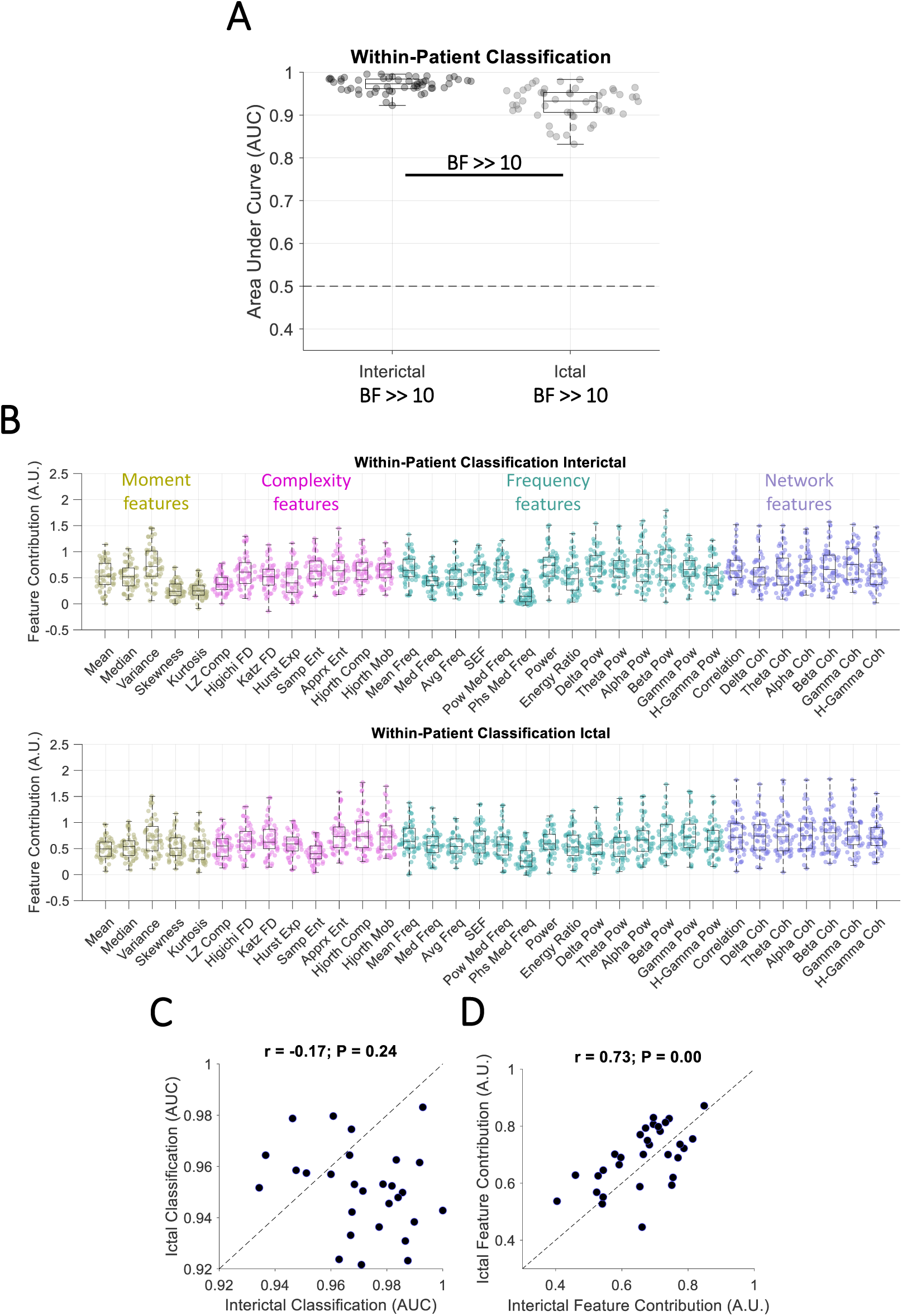
Classification of contacts with and without epileptogenic activities in each patient. **(A)** Area Under Curve (*AUC*) of classification performance for interictal and ictal classifications. Box plots show the distribution of data, its quartiles and median and whiskers indicate the maximum and minimum of the data over patients. Each dot indicates the data from one patient. Numbers below the bars indicate Bayesian evidence for the difference between true and null classification performances. Horizontal dashed line refers to theoretical chance-level classification (0.5). Bayes factor reflecting evidence for the difference between interictal and ictal classifications are also shown. *BF* >> 10 indicates *BF* > 100. **(B)** Contribution of each feature to the classification performance shown in A, calculated using random permutation. Yellow, pink, green and purple dots indicate moment, complexity, frequency and network-based features. **(C)** *Pearson* correlation between interictal and ictal classification performances across patients with each dot showing one patient. **(D)** *Pearson* correlation between interictal and ictal feature contributions across features with each dot showing one feature. Correlation and the corresponding *p* values are shown on top of panel **C** and **D** with the dashed line showing the theoretical perfect positive correlation.

Interestingly, there was significant evidence (*BF* >> 10) for higher classification in the interictal than ictal time window (Fig. 1A). This points to the importance of interictal neural activities in localisation of EZ. While the classification performances were high across all patients (*AUC* > 0.8), there was no correlation (*r =* −0. 17, *p* = 0.24; *Pearson*) between the level of *AUCs* in interictal and ictal time windows across patients (Fig. 1C). This suggests that patients with the clearest separation between epileptogenic and non-epileptogenic contacts in interictal window did not necessarily show the clearest separation between those contacts in the ictal window and vice versa.

We then evaluated the contribution of each feature to the performance (Fig. 1B). In interictal data, *variance* from the moment features, *Hjorth mobility* from the complexity features, *beta-band powe*r from the frequency features, and *gamma-band coherence* from the network features were among the most contributory features. In ictal data, *variance* from the moment features, *approximate entropy* from the complexity features, *gamma-band power* from the frequency features, and *beta-band coherence* from the network features were among the most contributory features. There was significant correlation between the features’ contributions across the two windows (*r =* 0.73, *p*<0.0001; *Pearson*; Fig. 1D) suggesting that similar sets of features dominantly contributed to the EZ localisation across interictal and ictal time windows.

Next, we evaluated the effect of outcome, region of resection (EZ), pathology and recording type on the classification results in each window (Supplementary Fig. 1). In the interictal data, there was evidence (1/10 < *BF* < 1/3) against any effect of outcome, region of resection, pathology and recording type on classification results. In the ictal data, there was significant evidence (*BF* < 1/10) against any effect of outcome, evidence (3 < *BF* < 10) for an effect of region of resection, insufficient evidence (1/3 < *BF* < 3) for an effect of pathology and evidence (1/10 < *BF* < 1/3) against any effect of recording type on classification results (Supplementary Fig. 1). To check the direction of region of resection effect, we used Bayes-factor *t-test* which showed insufficient evidence (1 < *BF* < 3) for higher classification in patients where the epileptogenic zone/resection was in temporal than frontal and mesiotemporal area (Supplementary Fig. 1).

Our classifications used all signal features simultaneously. To check if any individual feature could predict the resection outcome, we performed a direct comparison (unpaired Bayes factor *t-test*) between feature contributions in patients who became seizure-free (Engel I) vs. not seizure-free (Engel II-IV) outcomes (Supplementary Fig. 2). In interictal data, there was evidence (3 < *BF* < 10) that signal *median* contributed to better EZ localisation in patients who became seizure-free vs. those who did not. However, as *median* was among the least contributory features overall (c.f., Fig. 1B), we prefer not to put too much weight on this result. In ictal data, there was insufficient evidence (1/3 < *BF* < 3) for any feature to predict resection outcome.

As our features relied on signal patterns which were relatively sustained, compared to transient patterns such as interictal spikes or HFOs, we wondered whether accurate classification was possible using even shorter time windows. To test this, we repeated the classifications using the earliest, the middle and the latest 2-second time window of data in interictal and ictal data separately. Interestingly, we found significant evidence (*BF* >> 10) for above-chance *AUC* in both interictal and ictal time, with significant evidence (*BF* >> 10) for higher classification in ictal than interictal data (Supplementary Fig. 3). This repeated the pattern observed when using all windows of data in interictal and ictal periods (c.f., Fig. 1A).

### Do epileptiform patterns generalise across time windows?

We showed that a correlated set of features contributed to EZ localisation in both interictal and ictal windows (Fig. 1C), which might point to shared neural mechanisms underlying signal patterns in both time windows. We wondered if we could localise the EZ in the ictal window based on patterns of interictal activities. To test this, we trained our classifiers on interictal data and tested them on ictal data for each patient separately. We observed that, while the performance was lower (mean *AUC* = 0.60, *std* = 0.1; Fig. 2A) than those obtained by training and testing the classifiers within each time window separately (c.f., Fig. 1A), there was still significant evidence for above-chance cross-time generalisation (interictal to ictal; *BF* >> 10).

**Figure 2.**
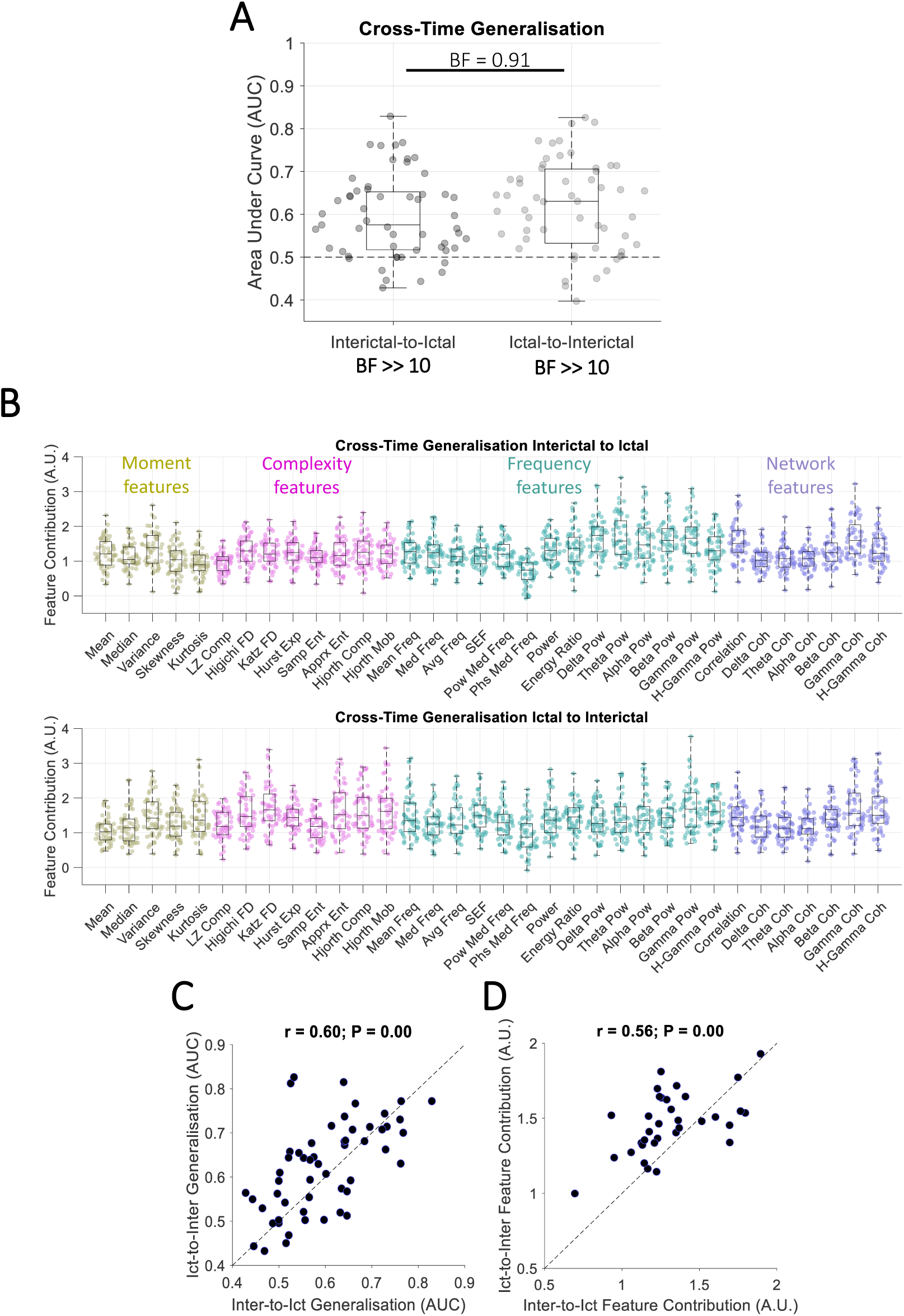
Classification of contacts with and without epileptogenic activities for each patient across time windows. **(A)** *AUC* of cross-time generalisation performance for interictal-to-ictal and ictal-to-interictal generalisations. Box plots show the distribution of data, its quartiles and median and whiskers indicate the maximum and minimum of the data over patients. Each dot indicates the data from one patient. Numbers below the bars indicate Bayesian evidence for the difference between true and null generalisation performances. Horizontal dashed line refers to theoretical chance-level generalisation (0.5). Bayes factor reflecting evidence for the difference between interictal-to-ictal and ictal-to-interictal generalisations are shown. *BF* >> 10 indicates *BF* > 100. **(B)** Contribution of each feature to the generalisation performance shown in A, calculated using random permutation. Yellow, pink, green and purple dots indicate moment, complexity, frequency and network-based features. **(C)** *Pearson* linear correlation between interictal-to-ictal and ictal-to-interictal generalisation performances across patients with each dot showing one patient. **(D)** *Pearson* linear correlation between interictal-to-ictal and ictal-to-interictal feature contributions across features with each dot showing one feature. Correlation and the corresponding *p* values are shown on top of panel **C** and **D** with the dashed line showing the theoretical perfect positive correlation.

Feature contribution results showed an advantage for *variance* from the moment features, *Higuchi fractal dimension* from the complexity features, *delta-band power* from the frequency features, and *gamma-band coherence* from network features (Fig. 2B). There was evidence (1/10 < *BF* < 1/3) against any effect of outcome, region of resection, pathology and insufficient evidence (1/3 < *BF* < 3) for an effect of recording type on the cross-time generalisation results (Supplementary Fig. 4).

Note that we only used two 5-minute windows of interictal recording to train the classifiers, which is relatively short given usual large clinical datasets of interictal activities recorded over several days. The generalisation performance is predicted to improve with higher volumes and more varied sets of training data. To check if increasing the diversity of the training data could improve the generalisation performance, we trained the classifiers using the ictal data and tested them using the interictal data. This would provide the classifiers with a more diverse training set as ictal data were obtained from more recordings than interictal recordings (3.7 vs. 2). Numbers of observations were equalised between interictal and ictal time windows.

While there was significant evidence (*BF* >> 10) for above-chance generalisation performance when training on the ictal data, there was insufficient evidence (*BF* = 0.91) for higher cross-time generalisation when the training data was from the ictal than interictal time windows. Therefore, while a more diverse dataset seems to have improved the classification (shifted the mean *AUC* from 0.6 up to 0.63), more data is needed to establish an improvement effect. There was significant correlation (*r =* 0.60, *p* < 0.001, *Pearson*; Fig. 2C) between the level of performance in interictal-to-ictal and ictal-to-interictal generalisations across patients. This shows that patients who showed the best generalisation from interictal to ictal windows also showed the highest generalisation in the opposite direction. This suggests that each patient has a certain level of similarity between interictal and ictal epileptiform patterns. There was significant correlation between the features’ contributions across the interictal-to-ictal and ictal-to-interictal generalisations (*r =* 0.56, *p* < 0.001; *Pearson*; Fig. 2D) suggesting that generalisable epileptiform patterns were reflected in similar sets of features no matter if generalising from interictal to ictal or vice versa.

In interictal-to-ictal generalisation data, there was evidence or significant evidence (*BF* > 3) that features of *Katz fractal dimension*, *energy ratio*, *theta-band power* led to poorer EZ localisation in patients with seizure-free vs. not seizure-free outcome (Supplementary Fig. 5). In ictal-to-interictal generalisation, this pattern was repeated for features of *energy ratio* and *delta-band power*. These suggest that specific features such as *energy ratio* might be more informative for EZ localisation when they show differences between their interictal and ictal patterns (i.e., as reflected in lower cross-time generalisability; Fig. 2B). This might mean that, patients whose signals’ *energy ratio* changes from interictal to ictal windows (e.g., through a significant increase) have a higher chance for their EZ to be localised; energy ratio change was indeed the basis for the original Epileptogenicity Index method^13^.

### Do epileptiform patterns generalise across patients?

So far, our analyses focused on within-time classification of contacts and cross-time generalisation of classifications both done within each patient. A clinically important aspect is to ascertain the generalisability of epileptiform patterns across patients, and test the feasibility of using the data from previous patients to help localise the EZ in new out-of-sample patients. To test this, we trained classifiers on the data from all patients minus one and tested the classifiers on the data from the left-out patient. This was done separately for interictal and ictal windows.

In interictal data, there was significant evidence (*BF* >> 10) for above chance cross-patient generalisation (Fig. 3A), which suggests that there were interictal epileptiform patterns which had similarities across patients. We evaluated the features’ contribution to the generalisation (Fig. 3B). Results showed an advantage for *kurtosis* from the moment features, *Higuchi fractal dimension* from the complexity features, *beta-band power* from the frequency features, and *gamma-band coherence* from the network features. There was significant evidence (*BF* < 1/10) against any effect of outcome, evidence (1/10 < *BF* < 1/3) against any effect of region of resection, insufficient evidence (1/3 < *BF* < 3) for an effect of pathology and significant evidence (*BF* > 10) for an effect of recording type on the cross-patient generalisation results (Supplementary Fig. 6). There was significant evidence (*BF* = 12) for higher generalisation to test patients with resection in mesiotemporal than temporal and significant evidence (*BF* >> 10) for higher generalisation to test patients with SEEG than ECoG recording (Supplementary Fig. 6).

**Figure 3.**
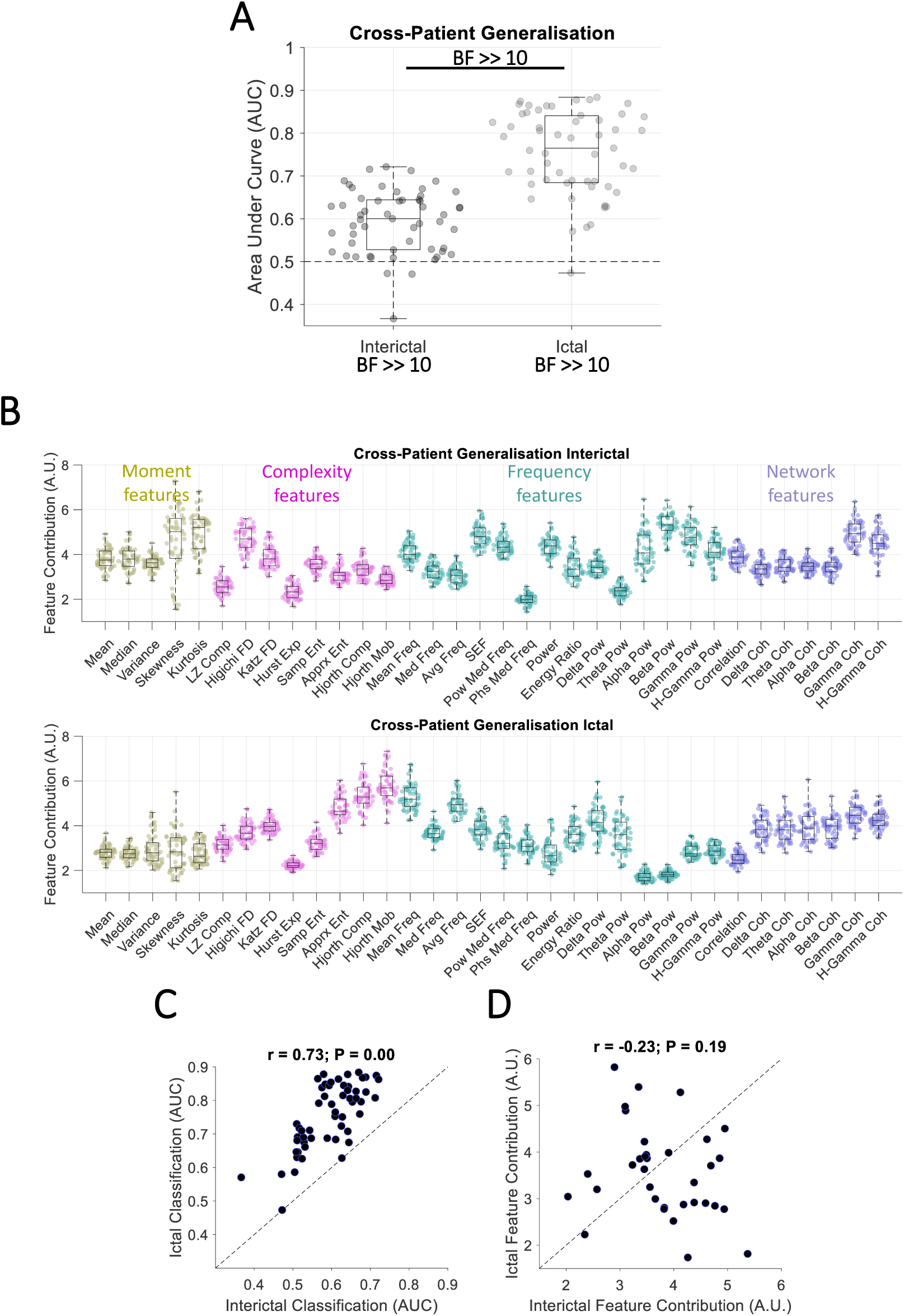
Classification of contacts with and without epileptogenic activities across patients. **(A)** *AUC* of cross-patient generalisation performance for interictal and ictal generalisations. Box plots show the distribution of data, its quartiles and median and whiskers indicate the maximum and minimum of the data over patients. Each dot indicates the data from one patient. Numbers below the bars indicate Bayesian evidence for the difference between true and null generalisation performances. Horizontal dashed line refers to theoretical chance-level generalisation (0.5). Bayes factor reflecting evidence for the difference between interictal and ictal generalisations are shown. *BF* >> 10 indicates *BF* > 100. **(B)** Contribution of each feature to the generalisation performance shown in A, calculated using random permutation. Yellow, pink, green and purple dots indicate moment, complexity, frequency and network-based features. **(C)** *Pearson* linear correlation between interictal and ictal generalisation performances across patients with each dot showing one patient. **(D)** *Pearson* linear correlation between interictal-to-ictal and ictal-to-interictal feature contributions across features with each dot showing one feature. Correlation and the corresponding *p* values are shown on top of panel **C** and **D** with the dashed line showing the theoretical perfect positive correlation.

In ictal data, there was significant evidence (*BF* >> 10) for above-chance cross-patient generalisation (Fig. 3A) which suggests that there were ictal epileptiform patterns which had similarities across patients. Results showed the highest contribution to generalisation by *skewness* from the moment features (however very low compared to other features), *Hjorth mobility* from the complexity features, *mean frequency* from the frequency features, and *gamma-band coherence* from the network features. There was evidence (1/10 < *BF* < 1/3) against any effect of outcome, insufficient evidence (1/3 < *BF* < 3) for an effect of region of resection and pathology and significant evidence (*BF* > 10) for an effect of recording type on the cross-patient generalisation. There was significant evidence (*BF* >> 10) for higher generalisation to test patients with resection in mesiotemporal than temporal and significant evidence (*BF* >> 10) for higher generalisation to test patients with SEEG than ECoG recording (Supplementary Fig. 6).

There was significant correlation (*r =* 0.73, *p* < 0.001; *Pearson*) between the level of generalisation in interictal and ictal time windows across patients (Fig. 3C). This suggests that if a patient’s data (i.e., testing set) has similarities to the pool of other patients’ data (i.e., training set), this will reflect in both interictal and ictal generalisations. On the other hand, patients with very distinct epileptiform activity patterns show this distinction across both interictal and ictal time windows. There was no correlation (*r =* −0.23, *p* = 0.19; *Pearson*) between informative features in the interictal and ictal time windows. This suggests that distinct sets of features contributed to cross-patient generalisation in interictal and ictal time windows.

In interictal data, there was significant evidence (*BF* >> 10; Bayes factor *t-test*) that *approximate entropy* led to better EZ localisation in patients who became seizure-free vs. those who did not (Supplementary Fig. 7). In ictal data, there was evidence or significant evidence (*BF* > 3) that *mean* and *median* led to better EZ localisation in patients who became seizure-free vs. those who did not and *median frequency* led to the opposite pattern. As these were all among the least contributory features to the overall cross-patient generalisation performance we avoid over-interpreting them.

## Discussion

Precise localisation of epileptogenic zone has remained a challenging problem. It has recently been highlighted that data and code sharing are fundamental to moving computational epilepsy studies towards clinical translation^11^. The current work uses one of the few open-access SEEG/ECoG datasets which provides epochs of interictal and ictal activities with meticulous labelling of electrode contacts, resection volume and clinical information including surgical outcome^46,48^. Using a rigorous ML-based pipeline, our study shows the feasibility of establishing generalisability of patterns within individuals from interictal to ictal periods, and across individuals during both interictal and ictal time windows. These results make several contributions to EZ localisation, explained below.

First, to establish that our ML-based method could discriminate areas with and without epileptiform activities, we classified epileptogenic and non-epileptogenic areas (EZ localisation, based on clinician labelling of the dataset) within each patient and found significant differences between the two classes of areas. At the individual patient level, we observed a remarkable EZ localisation performance in the interictal time window, which counter-intuitively surpassed that obtained in the ictal time window. Therefore, while majority of available studies have been developed to localise EZ during the ictal time window^13–19^, this study finds significant information within the interictal signal that can be utilised for EZ localisation. It is important to note that our approach did not pre-select any specific feature (such as spikes, HFO) as biomarkers *a priori*, but rather analysed the ensemble of the neural signal over time, which appears to contain predictive information beyond these well-known features, even when analysing time-windows as short as 2 seconds (c.f., Supplementary Fig. 3). This agrees with reports showing that interictal patterns are relatively stable over time^36,40,64^ (also see^65,66^).

While systematic comparison between interictal and ictal epileptiform patterns are rare in the literature, our result aligns with a surface EEG study in children with MRI-visible lesions which found better predictive value of interictal rather than ictal data^67^. We observed that the most informative features in interictal data included *variance*, *beta-band power*, *correlation* and *gamma-band coherence*, which in order support previous studies finding information in multiscale entropy in the gamma band^28,30^ (a complexity measure) and relative entropy^34^(a network measure). Interictal network studies have shown a gradient of within-area connectivity decreasing progressively from epileptogenic regions to propagation regions to non-involved zones^39,41,54^, providing a proxy for EZ localisation. The prominence of network measures in all of our analyses also aligns with a recent study showing greater information in network measures compared to univariate power-based measures in EZ localisation^48^. While we cannot perfectly equalise the interictal and ictal data for fair comparison as there are systematic differences in their collection time, number of epochs and potential artefacts in ictal signals, these results provide valuable insights into the richness of interictal activity patterns for EZ localisation.

The remarkable classification performance observed especially in the interictal window (> 0.90; Fig. 1A) supports the value of ML-based interictal localisation methods, which here used multiple features. While these features could have overlapped in their selectivity, they worked in synergy to detect as much non-overlapping information as possible. DT classifiers are good at combining distinct combinations of features to generate representational spaces in which classes can be separated. We also tested Support Vector Machine and Linear Discriminant Analysis classifiers, but both provided poorer classification. Our approach of using a range of simple-to-complex features is different from older localisation methods which used one main feature such as high-to-low frequency energy ratio^13,16^, and aligns with later studies which have combined several features for EZ localisation^17,31,68,69^, temporal detection of seizures^70^ and quantification of seizure severity^71^.

Second, we found prediction power in activity patterns of interictal data to localise EZ in the ictal period. While previous studies have localised EZ in interictal and ictal windows, the interictal-ictal correspondence has not been systematically investigated. In eleven patients with epilepsy who had been implanted with ECoG, one study found that the template of connectivity-based ictal epileptogenic areas could be helpful in informing the localisation of EZ interictally^72^. One recent study investigated the fine-grained timing and direction of interictal and ictal discharges using microelectrode grids, and suggested that interictal discharges are traveling waves that traverse the same path as seizure discharges^73^. After confirming a consistent temporal ordering of discharges in interictal and ictal windows, another study developed a novel source localisation method based on wave propagation, which successfully localised the EZ^74^. Our work evaluates the generalisability of a large battery of epileptiform features from interictal to ictal time windows and vice versa. The drop in cross-time generalisation performances (Fig. 2A) compared to the non-generalisation analysis (c.f., Fig. 1A) is supported by known differences in visualisable epileptogenic patterns from the interictal to the ictal time window^75,76^. In our study, the observed above-chance cross-time generalisation had not been necessarily predictable. It could be the case that epileptogenic areas would show higher value of a particular feature (e.g., power) than non-epileptogenic areas in the interictal window with this pattern flipping in the ictal window. This would have been detectable by our machine learning pipeline and would have reflected in below-chance (AUC < 0.5) generalisation performance. The potential of using interictal recording to predict EZ localisation is significant because many patients have insufficient or sometimes no seizures during their one/two-week hospitalisation for EZ localisation. This is an important limitation for visual localisation of EZ based on electrical patterns during seizures and for training ML algorithms, which, like humans, need enough samples to learn and localise epileptiform patterns from the data.

The third contribution of this work is showing that, despite clear inter-subject differences, there were patterns of epileptiform activities which were shared across patients. Machine learning allowed us to train the classifiers using data from one set of patients and test the generalisability of patterns to the data from a new out-of-sample patient. While these performances were expectedly lower in the cross-patient generalisation than within-patient classification (Fig. 3A vs. Fig. 1A), this result is promising and informative. The decrease in performance can be explained by large differences across patients’ data including epilepsy characteristics as well as distinct sampling of the brain, recording type, etc. Moreover, significant inter-subject differences may be present in terms of patient-specific epileptogenic “signatures”, the features of which are detectable across both interictal and ictal time windows for that individual^77^. On the other hand, a few studies showed that specific patterns can be generalised across patients, but only evaluated it in either interictal or ictal time window (Supplementary Table 1). A universal repertoire of seizure patterns across species has previously been observed, which suggests that some invariant properties characterise seizures under different physiological and pathological conditions^78^. Here, we showed that *ictal* epileptogenic patterns, especially those captured by complexity features, were more generalisable across subjects than *interictal* patterns. The present results also showed that SEEG recordings provided advantageous generalisability compared to ECoG (Supplementary Fig. 6). This is likely explained by SEEG’s greater sampling of a wider range of brain structures, which contributes to a more consistent sampling of the brain across patients^79^. These cross-patient generalisable patterns make it possible and desirable to use them on new out-of-sample data, to potentially build on these results by testing larger datasets. To facilitate future testing in novel datasets, we have shared our Matlab scripts.

One of the main concerns when using artificial intelligence in applications such as EZ localisation is the explainability of algorithms. Lack of knowledge about how a ML algorithm decides why a contact is classified as “epileptogenic” makes the algorithm less trustworthy for clinicians^80^, who may not be able to validate if a specific feature of a signal is indicative of epileptogenicity or whether the algorithm is simply wrong. Methodologies incorporating explainable features can mitigate the explainability issue and provide complementary insights into the growing body of work in EZ localisation and seizure prediction, which tend to adopt unexplainable ML algorithms such as deep neural networks^81,82^. Every individual feature used in our work has clear mathematical definition and has been validated in previous quantification analysis of neural data^55,56,83,84^. We also quantified the contribution of each feature in our analyses, thus avoiding the “black box” effect encountered when using algorithms such as deep neural networks. Accordingly, our proposed pipeline can be added as a primary feature extractor to prediction pipelines to make them more explainable to humans. It is of note that, while the mathematical definition of each of our features are clear, the neurophysiological correlates of these features needs to be sought for in the future.

There are several future directions which can facilitate the translation of this work to clinical practice. One can come through the improvement of the classification and generalisation performance. We used relatively short time windows of interictal (5-minute windows) and ictal (1-minute windows) data, both of which can be lengthened to potentially improve the classification performance. We did not apply any filtering or artefact removal, as ML classification algorithms are mainly sensitive to distinct patterns between classes (i.e., contacts with and without epileptiform patterns) rather than patterns which are common between classes (e.g., line noise). Nonetheless, one future direction would be to test if application of filters or artefact removal algorithms in the pre-processing stage can improve classification performance. Rather than an optimised work, the current study was only a feasibility effort to establish the generalisability of patterns of activities across time within patients and within time across patients. Another future direction would be to test the generalisability of the classification pipeline to datasets from other centres, which have undergone meticulous clinical evaluation and labelling. Finally, it would be interesting to evaluate the generalisability of the proposed pipeline to non-invasive modalities such as scalp EEG and magnetoencephalography (MEG). As the features extracted here are not modality-specific and rely on characteristics of time series, these methods can be applied to sensor- or source-space E/MEG data. The generalisability of these methods to non-invasive modalities is significant because current gold-standard invasive methods of SEEG and ECoG suffer from incomplete or sub-optimal sampling of the brain, and in addition these invasive methods are only indicated in a subset of patients. Delineation of likely spatial extent of epileptogenic zones could in theory be optimised using rigorous localisation methods developed here in conjunction with non-invasive E/MEG recording modalities, especially in the interictal time window. This will provide a more objective and fully automatic method for the localisation of EZ than current methods which often rely on visual detection, manual annotation and operator-dependent analysis of epileptiform patterns^85,86^.

In conclusion, we showed that powerful classification patterns were embedded within the EEG signal, which could reliably differentiate epileptogenic from non-epileptogenic contacts in every individual. Such patterns could be identified in both interictal and ictal recordings through features such as signal *variance*, *Hjorth mobility* and *complexity* as well as high-frequency power and network features, without taking account of any predetermined figures such as spikes, HFO or known ictal patterns. There were also features that could correctly predict EZ in ictal recordings from interictal recordings. Again, high-frequency *power* and network features were the most contributory features here. Finally, we showed that, while there were differences between epileptiform patterns across patients suggesting subject-specific effects, we could localise the EZ with well above chance precision using interictal and more dominantly ictal activities. The proposed methods and results provide new evidence for generalisability of epileptiform patterns across time and patients and open new avenues for future methods developed for epileptogenic zone localisation. Clues from neural signal changes could also provide new directions for investigating the biological correlates of interictal^87^ and ictal^88^ epileptiform activity. Their explainable nature is important for further investigation of pathophysiologic underpinnings of these signal changes. This could help contribute to efforts to develop paradigm-shifting therapeutic possibilities in epilepsy including disease-modifying treatments^89^, as well as further refining network-based surgical treatments^90^, seizure forecasting^91^ and seizure detection^92^.

## Data Availability

The dataset used in this study was from previous studies and is available at https://openneuro.org/datasets/ds004100/versions/1.1.3

## Footnotes

1 In this manuscript, the word “generalisation” refers to testing machine learning classifiers on data from unseen time windows/patients rather than the conventional epilepsy definition of seizure propagation over the brain.

## Acknowledgements

We would like to thank Mater Research Institute for supporting this study and Professor Brian Litt’s group for sharing their dataset online which allowed us to test our methods on their data.

## Funding

No funding was received towards this work.

## Competing interests

The authors report no competing interests.

## Notes

### Competing Interest Statement

The authors have declared no competing interest.

### Author Declarations

The study used (or will use) ONLY openly available human data that were originally located at: https://openneuro.org/datasets/ds004100/versions/1.1.3

### Summary of Updates

Correction of a few figure captions and addition of a few clarification statements to the manuscript.

