## Supplementary material for "Generalisability of epileptiform patterns across time and patients"

**Supplementary table 1. Overview of methods developed for EZ localisation.** The list presented is not comprehensive and represents the trend in method development for EZ localisation. Gen: cross-patient generalisation; Feature Category: which of this study's feature categories, would the methods fall into.

| Year | First author (Journal) | Gen. (Y/N) | Ictal/ Interictal | Category of features | Method description |
|---|---|---|---|---|---|
| 2008 | Bartolomei (Brain) | N | Ictal | Frequency | The ratio of high (e.g., >12 Hz) to low (e.g., <12 Hz) frequency energy and the time it significantly surpasses a threshold is combined into a single Epileptogenicity Index (EI). Channels with the highest EIs are defined as the Epileptogenic Zone (EZ). Threshold and frequencies are set manually. |
| 2011 | David (Brain) | N | Ictal | Frequency | Ratio of high (e.g., >12 Hz) to low (e.g., <12 Hz) frequency energy post seizure onset is statically compared to baseline (pre-onset) on cortical surface to localise EZ. Frequency bands are set manually. |
| 2014 | Gnatkovsky (Epilepsia) | N | Ictal | Moment & Frequency | Three markers of (i) power in high-(gamma) band activity, (ii) slow polarising shift and (iii) signal flattening are combined and a thresholding method defines the EZ. |
| 2018 | Grinenko (Brain) | Y | Ictal | Frequency | Three signal characteristics of (i) sharp transients or spikes, preceding seizure onset followed by (ii) multiband fast activity concurrent, with (iii) suppression of lower frequencies are combined using machine learning methods to localise EZ. |
| 2020 | Li (Hum Brain Mapp) | Y | | | |
| 2018 | Li (Netw Neurosci) | Y | Ictal | Network | Cross-power spectra in the gamma band (>30Hz) quantifies the connection between contacts and contact centrality trends over time localises EZ. |
| 2019 | Kini (Brain) | N | Ictal | Network | Inter-channel coherence (frequency-domain correlation) in different frequency bands (> 5 Hz) and broad-band time-domain correlation is calculated to develop a connectivity graph. The level of synchronisability indicates the EZ using manual thresholding. |
| 2020 | Balatskaya (Clin Neurophysiol) | N | Ictal | Frequency & Network | The original EI (see above) is combined with inter-contact directed correlation (out-degree). Thresholding localises EZ. |
| 2021 | Li (Nat Neurosci) | Y | Ictal | Network | Level of connectivity between contacts is quantified so that it keeps the data-driven dynamical model stable. Then the level of perturbation to contact connections which could destabilise the model indicates the most "*fragile*" contacts (EZ). |
| 2022 | Nakatani (Brain Commun) | N | Ictal | Frequency | Sustained direct current shifts and high-frequency (>80Hz) oscillations are manually detected at the start of seizure and used to localise EZ. |
| 2023 | Runfola (Commun Nonlin Sci Num Simulat) | N | Ictal | Network | In a dynamical network modelling, contacts with (i) high correlation, (ii) increased variance, and (iii) decorrelation from other contacts in the network indicate the EZ. |
| 2009 | Dauwels (Annu Int Conf IEEE | N | Interictal | Network | Cross-correlation, phase synchrony, magnitude coherence and Granger causality are used to localise hyper-synchronous areas which correspond to EZ. |

| | | | | | |
|---|---|---|---|---|---|
| | Eng Med Biol Soc) | | | | |
| 2011 | Gazit (J Neurosci Methods) | N | Interictal | Complexity & Network | Frequency-entropy measure is developed to quantify the inter-contact connectivity using time-frequency information. |
| 2012 | Andrzejak (Phys Rev E) | N | Interictal | Complexity & Network | Nonlinear randomness (defined as unpredictability of future values using present values) of signals and nonlinear dependence is quantified across contacts. Contacts within the EZ have higher non-randomness and inter-dependence. |
| 2016 | Mooij (Clin Neurophysiol) | N* | Interictal | Frequency | Mean and standard deviation of wavelet entropy are calculated in high-frequency bands (>80 Hz) to quantify signal complexity, which indicates higher complexity inside than outside the EZ. |
| 2019 | Sato (Epilepsy Behav) | N | Interictal | Complexity & Frequency | Regularity of gamma band (30-80 Hz) oscillations is quantified using multiscale entropy, which indicates higher regularity inside than outside EZ. |
| 2019 | Cimbalnik (Clin Neurophysiol) | Y | Interictal | Moment, Complexity, Frequency & Network | Multiple HFO-, univariate- (linear) and connectivity-based features, most dominantly, relative entropy, which evaluates the randomness and spectral richness between two contacts, is extracted in different frequency bands (1-1000 Hz) to localise EZ. |
| 2023 | Travnicek (Epilepsia) | N | | | |
| 2019 | Shah (Neuroimage Clin) | N | Interictal | Frequency & Network | Inter-channel (time-domain) correlation and coherence are calculated in different frequency bands (>5 Hz) which indicate higher values inside than outside the resected zone. |
| 2020 | Mooij (Clin Neurophysiol) | Y** | Interictal | Moment & Frequency | Skewness of signal power, which potentially correlates with spikes and HFOs, in different frequency bands (> 1 Hz) indicate higher skewness inside than outside EZ. |
| 2022 | Jiang (Adv Science) | N | Interictal | Network | Directed transfer entropy is calculated for contacts within and outside of EZ (here seizure onset zone) which showed higher inward flow for EZ contacts which is then used with decision tree classifiers to localise EZ. |
| 2022 | Taylor (Brain) | Y | Interictal | Frequency | Signal power in different frequency bands (>1 Hz) are calculated in non-epileptogenic contacts to develop a normative brain power map. Then, it is used for EZ localisation in new patients. |
| 2022 | Bernabei (Brain) | Y | Interictal | Frequency & Network | In addition to signal power (see the above row), inter-channel coherence is quantified in different frequency bands (> 1 Hz) to develop a normative brain map. Then, it is used for EZ localisation in new patients. |
| 2022 | Gunnarsdottir (Brain) | Y | Interictal | Network | A dynamical system modelling algorithm is developed to determine contacts which dominantly send or receive signals. The receiving contacts (“sinks”) are dominantly inside the EZ. |
| 2023 | Diamond (Brain) | N | Interictal | Network | Interictal discharges are shown to follow the same direction as seizure waves. An algorithm uses time differences of discharge receipt at nearby electrodes to localise EZ. |

| | | | | | |
|---|---|---|---|---|---|
| 2023 | Johnson (Brain) | Y | Interictal | Network | Undirected and directed connectivity are quantified in different frequency bands (>4 Hz) using coherence and partial directed coherence, respectively. Connectivity is dominantly inward and higher for contacts inside than outside the EZ. |

* Data from all patients were concatenated and analysed simultaneously, and generalisation was not checked systematically.

** Data from several patients were concatenated separately for training and testing sets.

**Supplementary table 2** Demographics of patients.

| Age | Sex | Hand | Outcome | Engel | Therapy | Implant | Resection target | Lesion status | Onset age |
|---|---|---|---|---|---|---|---|---|---|
| 40-49 | F | R | F | 3A | ABLATION | SEEG | FRONTAL | n/a | 10-19 |
| 20-29 | M | L | S | 1D | RESECTION | ECOG | FRONTAL | LESIONAL | 0-9 |
| 30-39 | M | R | S | 1B | RESECTION | ECOG | TEMPORAL | LESIONAL | 0-9 |
| 30-39 | M | L | S | 1B | RESECTION | ECOG | FRONTOPARIETAL | NON-LESIONAL | 10-19 |
| 20-29 | F | L | S | 1C | RESECTION | ECOG | TEMPORAL | LESIONAL | 0-9 |
| 50-59 | F | L | F | 4A | RESECTION | ECOG | TEMPORAL | NON-LESIONAL | 50-59 |
| 40-49 | F | L | F | 2C | RESECTION | ECOG | TEMPORAL | NON-LESIONAL | 30-39 |
| 50-59 | F | R | S | 1A | RESECTION | ECOG | TEMPORAL | LESIONAL | 30-39 |
| 20-29 | F | L | F | 2A | RESECTION | ECOG | TEMPORAL | NON-LESIONAL | 10-19 |
| 20-29 | M | L | S | 1D | RESECTION | ECOG | FRONTAL | LESIONAL | 10-19 |
| 30-39 | F | L | S | 1D | RESECTION | ECOG | TEMPORAL | LESIONAL | 0-9 |
| 20-29 | M | n/a | S | 1B | RESECTION | ECOG | TEMPORAL | LESIONAL | 20-29 |
| 40-49 | F | R | S | 1B | RESECTION | ECOG | TEMPORAL | NON-LESIONAL | 20-29 |
| 30-39 | F | n/a | S | 1D | RESECTION | ECOG | TEMPORAL | NON-LESIONAL | 30-39 |
| 30-39 | M | R | S | 1A | RESECTION | ECOG | TEMPORAL | LESIONAL | 20-29 |
| 40-49 | F | L | S | 1B | RESECTION | ECOG | TEMPORAL | NON-LESIONAL | 20-29 |
| 30-39 | M | R | S | 1A | RESECTION | ECOG | TEMPORAL | NON-LESIONAL | 0-9 |
| 40-49 | F | R | S | 1B | RESECTION | ECOG | TEMPORAL | NON-LESIONAL | 20-29 |
| 20-29 | F | R | F | 3A | ABLATION | SEEG | FRONTAL | LESIONAL | 0-9 |
| 40-49 | F | n/a | F | 3A | ABLATION | ECOG | MESIOTEMPORAL | NON-LESIONAL | 20-29 |
| 50-59 | F | R | S | 1A | ABLATION | SEEG | MESIOTEMPORAL | LESIONAL | 40-49 |
| 30-39 | M | L | S | 1A | RESECTION | SEEG | TEMPORAL | LESIONAL | 10-19 |
| 30-39 | M | n/a | S | 1A | RESECTION | ECOG | TEMPORAL | LESIONAL | 30-39 |
| 20-29 | F | n/a | S | 1A | ABLATION | ECOG | MESIOTEMPORAL | NON-LESIONAL | 22 |
| 40-49 | F | L | S | 1B | ABLATION | SEEG | MFL | NON-LESIONAL | 20-29 |
| 50-59 | F | L | F | 3A | ABLATION | SEEG | MESIOTEMPORAL | NON-LESIONAL | 40-49 |
| 30-39 | M | n/a | S | 1B | RESECTION | SEEG | FRONTAL | LESIONAL | 0-9 |
| 30-39 | M | R | F | 2A | ABLATION | SEEG | MESIOTEMPORAL | NON-LESIONAL | 30-39 |
| 30-39 | M | L | F | 4A | ABLATION | SEEG | MESIOTEMPORAL | LESIONAL | 20-29 |
| 20-29 | M | L | S | 1A | ABLATION | SEEG | PARIETAL | LESIONAL | 0-9 |
| 40-49 | F | L | S | 1B | ABLATION | SEEG | MESIOTEMPORAL | NON-LESIONAL | 20-29 |
| 30-39 | M | R | S | 1C | ABLATION | SEEG | MESIOTEMPORAL | NON-LESIONAL | 10-19 |
| 30-39 | M | n/a | S | 1D | ABLATION | SEEG | MESIOTEMPORAL | LESIONAL | 10-19 |
| 30-39 | M | n/a | S | 1D | RESECTION | SEEG | TEMPORAL | LESIONAL | 0-9 |
| 10-19 | M | n/a | S | 1A | RESECTION | SEEG | TEMPORAL | NON-LESIONAL | 0-9 |
| 20-29 | M | n/a | S | 1A | ABLATION | SEEG | TEMPORAL | LESIONAL | 10-19 |

| | | | | | | | | | |
|---|---|---|---|---|---|---|---|---|---|
| 10-19 | M | R | S | 1B | ABLATION | SEEG | INSULAR | LESIONAL | 0-9 |
| 30-39 | M | R | F | 2A | ABLATION | SEEG | MFL | NON-LESIONAL | 0-9 |
| 30-39 | M | R | F | 3A | ABLATION | SEEG | INSULAR | NON-LESIONAL | 0-9 |
| 40-49 | F | n/a | S | 1A | RESECTION | SEEG | TEMPORAL | NON-LESIONAL | 10-19 |
| 30-39 | F | n/a | F | 3A | ABLATION | SEEG | MESIOTEMPORAL | NON-LESIONAL | 10-19 |
| 40-49 | F | L | S | 1D | ABLATION | SEEG | MESIOTEMPORAL | NON-LESIONAL | 10-19 |
| 30-39 | F | L | S | 1D | ABLATION | SEEG | MESIOTEMPORAL | LESIONAL | 10-19 |
| 20-29 | M | n/a | F | 3A | RESECTION | SEEG | TEMPORAL | LESIONAL | 0-9 |
| 50-59 | M | L | F | 2A | ABLATION | SEEG | FRONTAL | NON-LESIONAL | 0-9 |
| 20-29 | F | L | F | 2A | ABLATION | SEEG | FRONTAL | NON-LESIONAL | 0-9 |
| 20-29 | F | R | S | 1A | RESECTION | SEEG | TEMPORAL | LESIONAL | 10-19 |
| 40-49 | F | R | S | 1A | RESECTION | SEEG | TEMPORAL | NON-LESIONAL | 0-9 |
| 20-29 | F | L | F | 3A | RESECTION | SEEG | FRONTAL | LESIONAL | 10-19 |
| 20-29 | F | L | S | 1A | ABLATION | SEEG | FRONTAL | LESIONAL | 0-9 |
| 30-39 | F | L | F | 3A | ABLATION | SEEG | TEMPORAL | LESIONAL | 10-19 |
| 30-39 | M | L | S | 1A | ABLATION | SEEG | MESIOTEMPORAL | LESIONAL | 0-9 |
| 20-29 | M | n/a | F | 2A | ABLATION | SEEG | MESIOTEMPORAL | NON-LESIONAL | 10-19 |
| 20-29 | F | L | F | 3A | RESECTION | SEEG | FRONTAL | LESIONAL | 0-9 |
| 20-29 | M | L | F | 3A | RESECTION | SEEG | MESIOTEMPORAL | NON-LESIONAL | 10-19 |

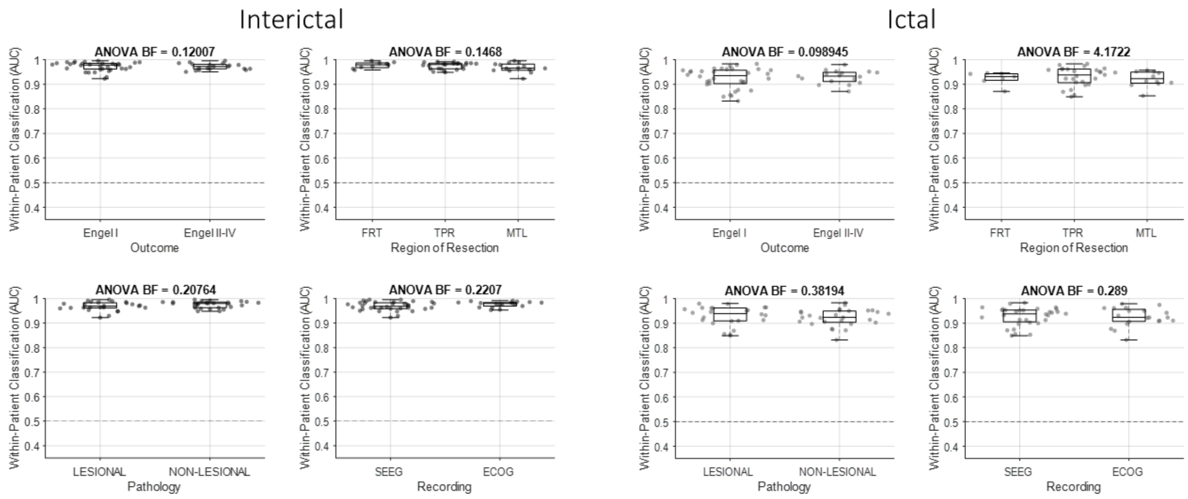


**Supplementary figure 1 Main effects of resection outcome, area of resection, pathology (lesion/non-lesion) and recording type (SEEG/ECoG) on EZ localisation.** Effects were calculated using Bayes factor *ANOVA*. Left and right panels show the results for interictal and ictal classification analysis, respectively. Box plots show the distribution of data, its quartiles and median and whiskers indicate the maximum and minimum of the data over patients. Each dot indicates the data from one patient. Horizontal dashed line refers to theoretical chance-level classification (0.5). The number on the top of each panel indicates Bayesian evidence for an effect on classification performance. Bayes factor *t-test* results reflecting evidence for the difference between pairs of conditions are shown only if BF > 3. BF >> 10 indicates BF > 100. FRT: frontal; TPR: temporal; MTL: mesiotemporal.

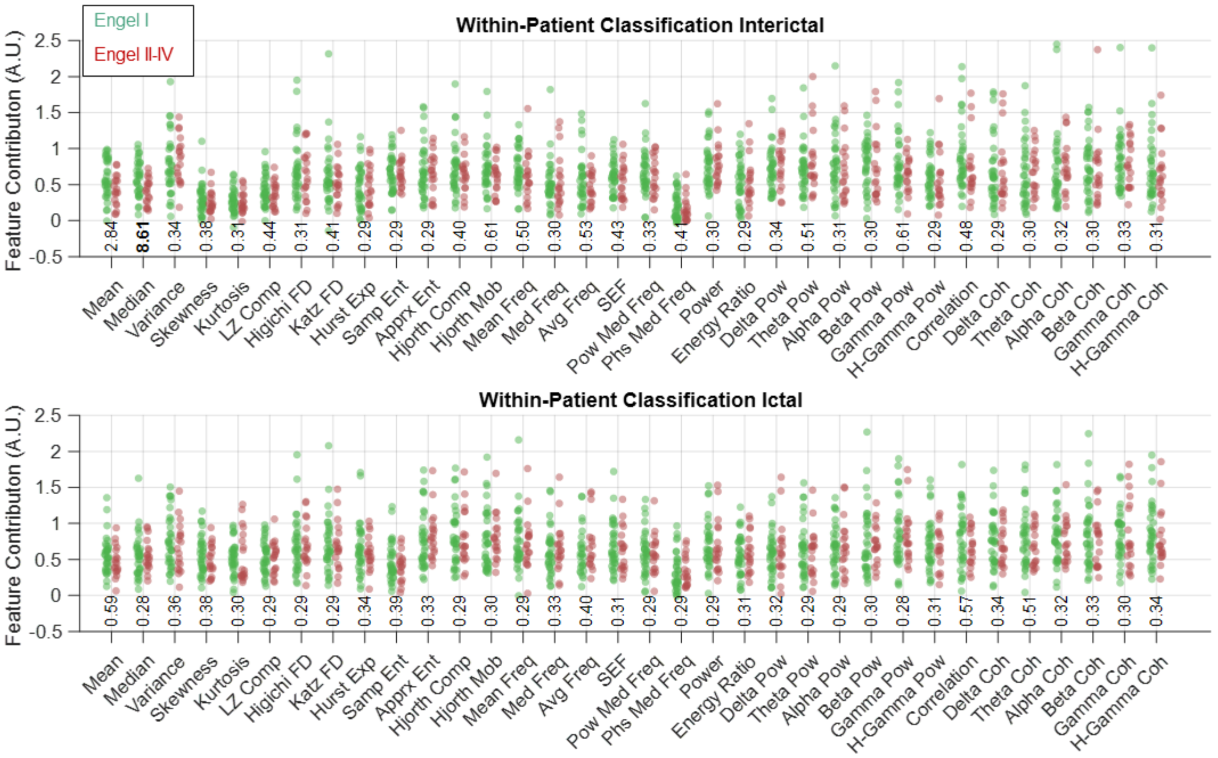


**Supplementary figure 2 Effect of resection outcome on feature contribution to classification of contacts with and without epileptiform activities.** Each dot indicates the data from one patient. Patients are grouped into those who became seizure-free (Engel I, green) and those who did not (Engel II-IV, red). Top and bottom panels show the results for interictal and ictal classification analysis, respectively. Number at the top of each feature indicates Bayesian *t-test* evidence for an effect of resection outcome on feature contribution, with bold fonts indicating evidence and significant evidence (BF >10) for difference between patients with and without seizure-free outcomes.

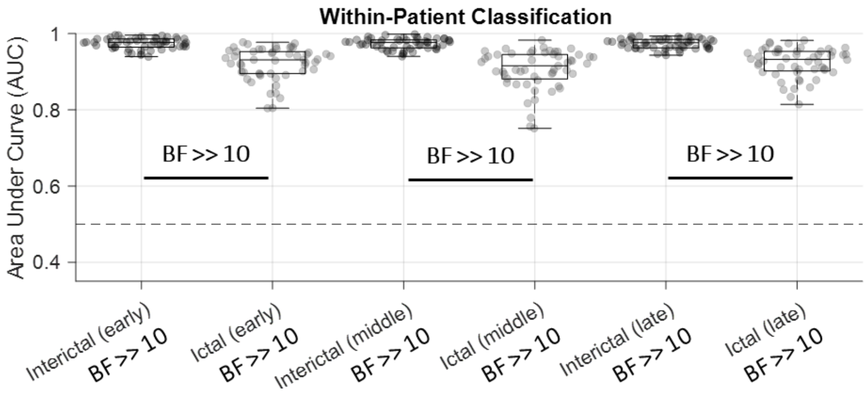


**Supplementary figure 3 Classification of contacts with and without epileptogenic activities across patients using an early, middle and late time window of data.** Area Under Curve (AUC) of classification performance for interictal and ictal classifications. Box plots show the distribution of data, its quartiles and median and whiskers indicate the maximum and minimum of the data over patients. Each dot indicates the data from one patient. Numbers below the bars indicate Bayesian evidence for the difference between true and null classification performances. Horizontal dashed line refers to theoretical chance-level classification (0.5). Bayes factor reflecting evidence for the difference between interictal and ictal classifications are also shown. BF >> 10 indicates BF > 100.

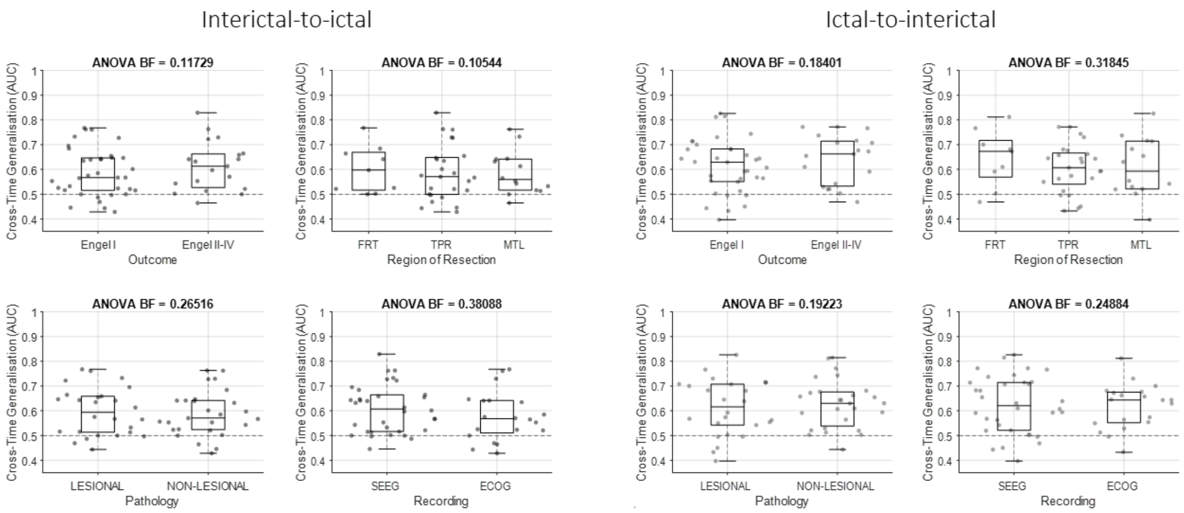


**Supplementary figure 4 Main effects of resection outcome, region of resection, pathology (lesion/non-lesion) and recording type (SEEG/ECoG) on generalisation of EZ localisation information across time.** Effects were calculated using Bayes factor *ANOVA*. Left and right panels show the results for interictal-to-ictal and ictal-to-interictal generalisation analysis, respectively. Box plots show the distribution of data, its quartiles and median and whiskers indicate the maximum and minimum of the data over patients. Each dot indicates the data from one patient. Horizontal dashed line refers to theoretical chance-level generalisation (0.5). The number on the top of each panel indicates Bayesian evidence for an effect on generalisation performance. Bayes factor *t-test* results reflecting evidence for the difference between pairs of conditions are shown only if BF > 3. BF >> 10 indicates BF > 100. FRT: frontal; TPR: temporal; MTL: mesiotemporal.

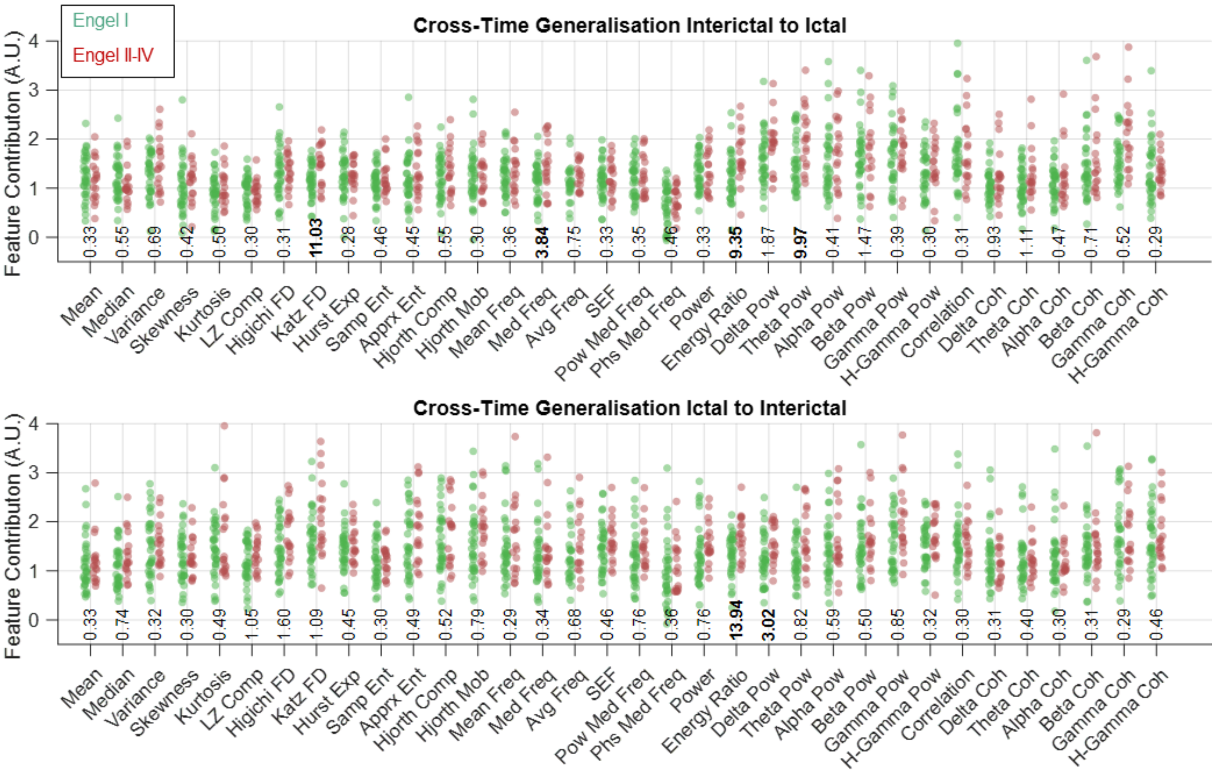


**Supplementary figure 5 Effect of resection outcome on feature contribution to classification of contacts with and without epileptiform activities.** Each dot indicates the data from one patient. Patients are grouped into those who became seizure-free (Engel I, green) and those who did not (Engel II-IV, red). Top and bottom panels show the results for interictal-to-ictal and ictal-to-interictal generalisation analysis, respectively. The number at the top of each feature indicates Bayesian *t-test* evidence for an effect of resection outcome on feature contribution, with bold fonts indicating evidence and significant evidence (BF >10) for difference between patients with and without seizure-free outcomes.

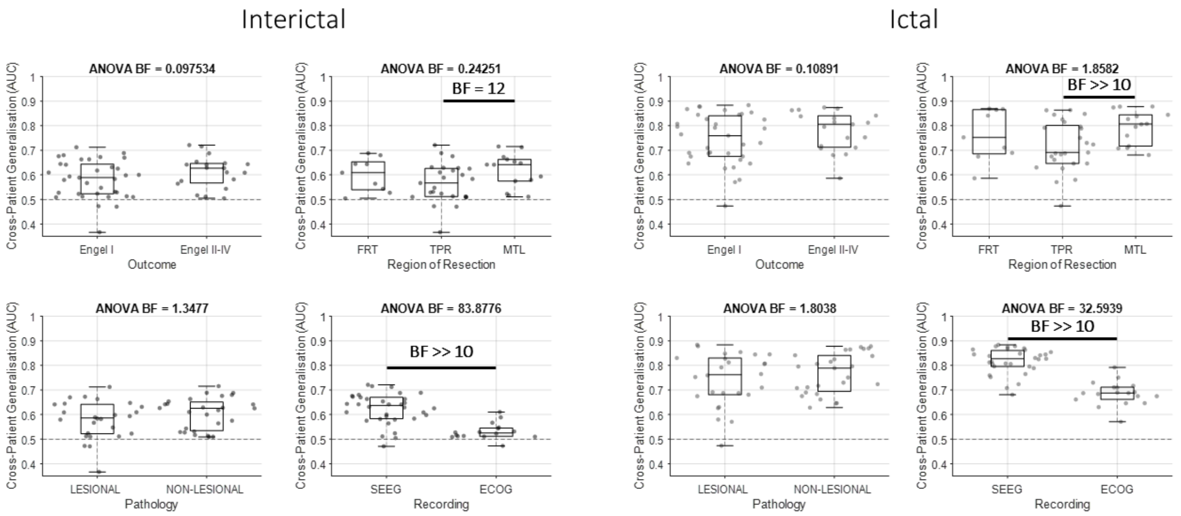


**Supplementary figure 6 Main effects of resection outcome, region of resection, pathology (lesion/non-lesion) and recording type (SEEG/ECoG) on generalisation of EZ localisation information across patients**. Effects were calculated using Bayes factor *ANOVA*. Left and right panels show the results for interictal and ictal generalisation analysis, respectively. Box plots show the distribution of data, its quartiles and median and whiskers indicate the maximum and minimum of the data over patients. Each dot indicates the data from one patient. Horizontal dashed line refers to theoretical chance-level generalisation (0.5). The number on the top of each panel indicates Bayesian evidence for an effect on generalisation performance. Bayes factor *t-test* results reflecting evidence for the difference between pairs of conditions are shown only if BF > 3. BF >> 10 indicates BF > 100. FRT: frontal; TPR: temporal; MTL: mesiotemporal.

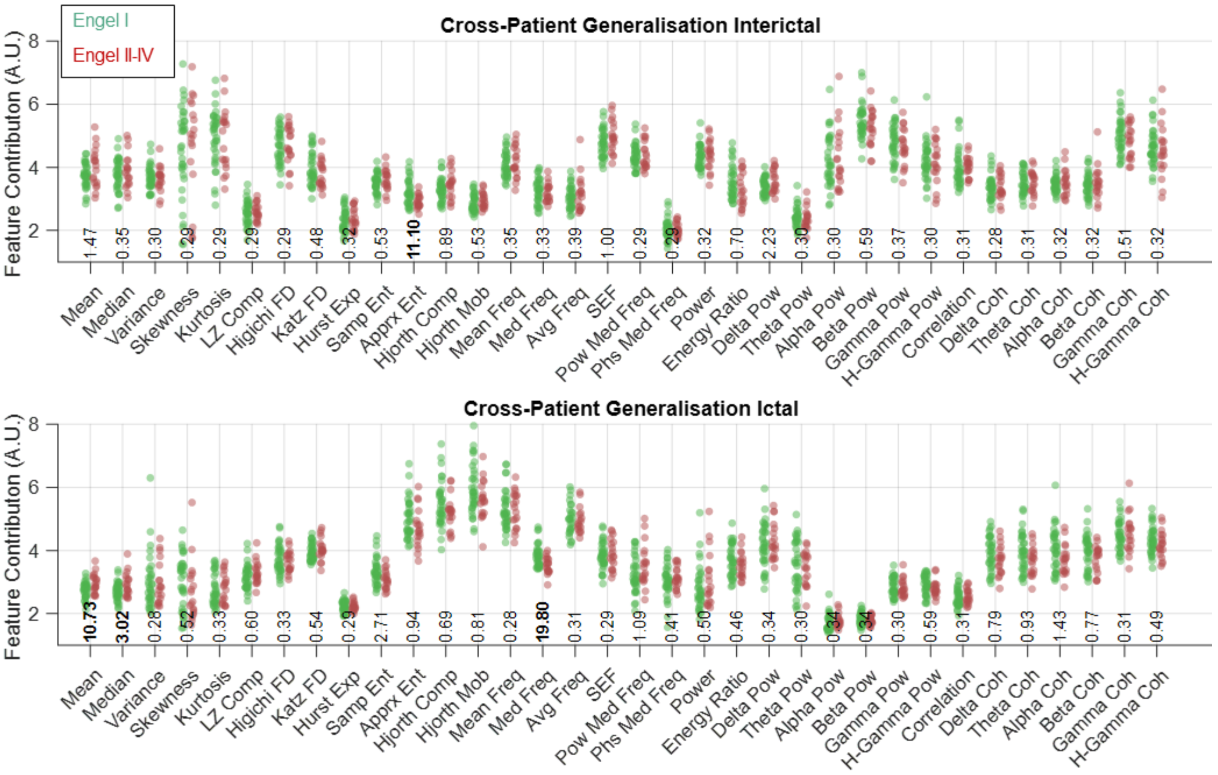


**Supplementary figure 7 Effect of resection outcome on feature contribution to classification of contacts with and without epileptiform activities.** Each dot indicates the data from one patient. Patients are grouped into those who became seizure-free (Engel I, green) and those who did not (Engel II-IV, red). Top and bottom panels show the results for interictal and ictal cross-patient generalisation analysis, respectively. Number at the top of each feature indicates Bayesian *t-test* evidence for an effect of resection outcome on feature contribution, with bold fonts indicating evidence and significant evidence (BF >10) for difference between patients with and without seizure-free outcomes.

### Supplementary text 1

Signal moment features included signal *Mean*, *Variance, Skewness* and *Kurtosis* which are the four statistical moments of time series. Signal *Mean* provides an estimate of average amplitude of samples within the time series and signal *Variance* quantifies their variability around the *Mean*. Therefore, the more scattered the samples, the higher the *Variance*. *Skewness* quantifies how deviated majority of signal samples are relative to their *Mean*. This leads to a high *Skewness* when majority of signal samples are away from the *Mean* vs. near the *Mean*. *Kurtosis* quantifies how densely concentrated signal samples are around their *Mean*. Therefore, *Kurtosis* is higher when a higher vs. a fewer number of signal samples are concentrated around the *Mean*. We also extracted signal Median as it can detect pattern differences which can be missed by *Mean*[1].

Signal nonlinear complexity features reflect how complex vs. predictable signal patterns are (see[2] for a review). We quantified the complexity of signals using 8 distinct complexity metrics. LZ Complexity (*LZ Comp*) quantifies the complexity of time series by counting the number of unique sub-sequences of samples in the time series after turning the signal into a binary sequence using a median-based threshold[3]. Accordingly, a sequence of samples with a certain regularity leads to a lower LZ complexity. Fractal dimensions provide statistical information determining the complexity of how data is organized within the signal[4,5]. Accordingly, a higher fractal value indicates more complexity for a time series as reflected in more nesting of repetitive sub-sequences at all scales. We extracted two common methods for quantifying fractal dimensions namely Higuchi and Katz fractal dimensions (*Higuchi and Katz FD*). Hurst Exponent (*Hurst Exp*) quantifies the long-term "memory" in signals by calculating the degree of dependence among its consecutive samples[6]. Therefore, it operates like an autocorrelation function over time. Sample and Approximate Entropies (*Samp* and *Apprx Ent*) are two methods for quantifying the level of perturbation in the signal[7]. Each of these entropy metrics have specific properties. Sample entropy is not as sensitive to the sample size and simpler to implement compared to approximate entropy. Sample entropy, however, does not consider self-similar patterns in the time series. Hjorth Complexity and Mobility (*Hjorth Comp* and *Mob*) measure the variation in signal characteristics[3]. The complexity measure calculates the variation in the signal's dominant frequency, and the Mobility measures the width of the signal power spectrum - how widely the frequencies are scattered in the power spectrum of the signal. A signal with a wider range of frequencies will lead to higher Hjorth complexity and mobility.

We also extracted 14 frequency-domain features from the signals using the Power Spectrum Density (PSD), to cover the features which recent studies also showed their value[8–10]. PSD represents the intensity or the distribution of the signal power into its constituent frequency components. This included Mean Frequency (*Mean Freq*) which is the average of all frequency components available in a signal. We also extracted Median Frequency (*Med Freq*), which is the median normalized frequency of the PSD, as well as signal Power and Phase at Median Frequency (*Pow Med Freq* and *Phs Med Freq*) as two additional features. Average Frequency (*Avg Freq*) was also quantified as the number of times the signal time series crosses zero. We extracted Spectral Edge Frequency (*SEF*) which indicates the highest frequency below which x percent of the signal's power spectrum exists. X was set to 95% in this study. Therefore, SEF reflects the upper-bound of frequency in the PSD. As a general indicator of epileptogenicity[11], signal Power (Power) was also extracted over the whole PSD as well as in delta (*Delta Pow*; 0 - 4 Hz), theta (*Theta Pow*; 4 - 8 Hz), alpha (*Alpha Pow*; 8 – 13 Hz), beta (*Beta Pow*; 13 – 30 Hz), gamma (*Gamma Pow*; 30 – 90 Hz) and high gamma (*H-Gamma Pow*; 90 Hz to sampling frequency/2 Hz). To be able to compare the results with energy-ratio-based metric of epileptogenicity, we also extracted *Energy Ratio* as the ratio of signal energy in the high (12.4 to 97 Hz) over the low (3.5 to 12.4 Hz) frequency bands[12]. For that purpose, signals were band-pass-filtered separately in the mentioned frequency bands and the ratio of signal energy in their time samples was used as *Energy Ratio*[13]. *Energy ratio* is the basic feature used in the epileptogenicity index which is higher upon seizure onset in epileptic vs. non-involved contacts when seizure causes high-frequency oscillations in the signals [12].

Finally, following the recent shift towards network-based analysis in epilepsy, we also extracted 7 features which quantified how connected a contact was to other contacts. Previous studies have shown that epileptic areas can be delineated from other areas through network/connectivity metrics[9,14–20.] To see if network measures could indicate EZ, we extracted inter-contact zero-lagged correlation (*Correlation*) as well as inter-contact coherence in delta (*Delta Coh*; 0 - 4 Hz), theta (*Theta Coh*; 4 - 8 Hz), alpha (*Alpha Coh*; 8 – 13 Hz), beta (*Beta Coh*; 13 – 30 Hz), gamma (*Gamma Coh*; 30 – 90 Hz) and high gamma (*H-Gamma Coh*; 90 Hz to sampling frequency/2 Hz), which have been recently used in epilepsy network analysis[8]. While inter-contact zero-lagged correlation provides one of simplest measures of inter-area connectivity in the time domain, coherence is a frequency-domain metric which uses each area's PSD and their cross-frequency connectivity. To obtain a single value of connectivity for each contact, we averaged all the connectivity values obtained between the given contact and all the other ones.
